# Weak relational ties and ineffective communication in vocational rehabilitation: a case study of people on long-term sick leave with common mental disorders

**DOI:** 10.1101/2025.07.03.25330864

**Authors:** Frederik Martiny, Marius Brostrøm Kousgaard, Christian Jauernik, Pia Kürstein Kjellberg, Maarten Rozing, Frans Waldorff, Mads Aage Toft Kristensen

## Abstract

2

**Purpose:** To explore how collaborative challenges are experienced and navigated among citizens on long-term sick leave due to common mental disorders and vocational rehabilitation professionals.

**Methods:** A qualitative case study in two Danish municipalities, including semi-structured interviews with seven citizens and seven GPs, and six focus group interviews with 31 CWs. Data were analysed inductively using systematic text condensation followed by abductive interpretation informed by relational coordination theory.

**Results:** Collaborative challenges among citizens and professionals were rooted in weak relational ties and reinforced by counterproductive communication. A lack of shared goals led to collaboration being experienced as a competition over the agenda, reinforced by campaigning communication aimed at persuading others of one’s perspective. A lack of shared knowledge made collaboration feel colossally laborious, reinforced by clarifying communication aimed at reducing ambiguity through repeated explanations. A lack of mutual trust and respect contributed to collaborators being perceived as adversaries, reinforced by self-protective communication aimed at shielding oneself from criticism.

**Conclusion:** Collaborative challenges in vocational rehabilitation can be understood as self-reinforcing cycles of weak relational ties, negative collaborative experiences, and counterproductive communication. Strengthening relational coordination may improve collaboration for people on long-term sick leave due to common mental disorders.

## 4 INTRODUCTION

Mental disorders are prevalent, affecting nearly one in seven people on a global scale, with a median age of onset of 18 years (1,2). In recent years, the prevalence of the most common mental disorders (common mental disorders), including anxiety, mild to moderate depression, adjustment disorders, and stress-related illnesses, has significantly increased, especially in the working age population (age 15-64 years) and among women who are disproportionately affected by common mental disorders compared to men (3). Living with common mental disorders has serious personal consequences, including reduced quality of life, disability, and stigma (4–6). Also, mental disorders place a significant burden on societies through healthcare costs, lost productivity, and social care expenses (3–8).

In terms of employment, the OECD estimates that common mental disorders account for 40% of sick leave and are the leading cause of absenteeism (5,9). In turn, people on sick leave due to common mental disorders have a three times lower likelihood of returning to work compared to other sick leave causes (5,10). Resultantly, people on sick leave due to common mental disorders are at risk of long-term unemployment, which is a strong negative social determinant of health, causing financial strain, social isolation, and poor health (11–13). In turn, unemployment and mental illness are associated with stigma, including feelings of shame and guilt for not contributing to the labour market, having a “lifestyle disease” or feeling like “a burden to society” (14–16). Stigma adds to the negative health effects of unemployment (6), contributing to vicious cycles between poor health, worsened social conditions and marginalisation from the labour market among unemployed people with common mental disorders (5,13,17–21). At the societal level, these self-reinforcing vicious cycles contribute to widening health inequities, as socioeconomically disadvantaged groups face a higher risk of developing common mental disorders and experiencing prolonged exclusion from the labour market (5,17,22).

Therefore, providing employment, health, and social care support to unemployed people with health-related concerns is a public health priority (23). Such support is commonly referred to as vocational rehabilitation, aiming to improve work participation among working-age individuals whose health affects their ability to work. Of note, vocational rehabilitation requires collaboration among multiple stakeholders across sectors, institutions, and professional disciplines (23–28). Consequently, its effectiveness depends on the quality of collaboration between stakeholders (5,25,27,28).

Unfortunately, many barriers impede effective collaboration in vocational rehabilitation. On a societal level, differing legal frameworks can hinder collaboration, as can incongruent IT systems, shifting political goals, and conflicting paradigms among professional institutions (25–27,29–31). At the organisational level, staff turnover, time constraints, reliance on written communication, incompatible work procedures, latent distrust, and uncertainties about roles and responsibilities across institutions can limit professionals’ ability to coordinate their tasks effectively (24–27,31–36). These macro-level barriers lead to interpersonal challenges among stakeholders in vocational rehabilitation. Studies of collaboration experiences in vocational rehabilitation among professionals report struggles with misalignment of mutual expectations, difficulty establishing shared goals, a lack of a shared knowledge base, and disagreements about how to communicate effectively with each other (24,26,27,31,34–37). Studies of citizens’ experiences of the collaboration have found uncertainty about the return-to-work process and expectations regarding task delegation among stakeholders, difficulties in establishing trust and in feeling involved and validated in interactions with professionals, and fragmented or poorly coordinated services (24,25,27–30,32–34,36).

Collaborative challenges in vocational rehabilitation among professionals and citizens have proven quite similar over time, across target groups, and across countries (26–28,35,38–40), despite substantial differences in the organisation of vocational rehabilitation services (5,38). This consistency across different contexts suggests that deeper-rooted interpersonal dynamics might both cause and perpetuate collaborative challenges in vocational rehabilitation for people on sick leave. However, to our knowledge, previous research in the field of vocational rehabilitation has not studied how the interpersonal dynamics between professionals and citizens contribute to, and potentially sustain, collaborative challenges among them.

Therefore, with this study, we aimed to explore how collaborative challenges are experienced and navigated among citizens on long-term sick leave due to common mental disorders and vocational rehabilitation professionals.

## 5 METHODS

### 5.1 METHODOLOGY

We conducted a case study using qualitative methods in two municipal settings in the Capital Region of Denmark, including the perspectives of citizens, CWs, and GPs concerning collaborative challenges in vocational rehabilitation for citizens on long-term sick leave due to common mental disorders. Data generation for the study was conducted between December 2022 and May 2023.

### 5.2 SETTING

#### 5.2.1 THE DANISH SICK LEAVE SYSTEM

In Denmark, vocational rehabilitation for people on sick leave is delivered by various providers working across the health, social, and employment sectors. Denmark is divided into five regions responsible for providing healthcare services, including both primary and secondary care. Municipalities are responsible for social, preventive health, and employment services. Denmark’s 98 municipalities are legally required to deliver employment services through a job centre, which is commonly organised separately from their preventive health and social services (39). The municipal job centres employ CWs who coordinate the vocational rehabilitation process and support citizens throughout their sick leave. Most CWs hold a professional bachelor’s degree in social work. However, it is not uncommon for CWs to have other educational backgrounds and to acquire the necessary competencies through on-the-job training.

Sickness benefits are provided to people on sick leave for up to 22 weeks, contingent on their participation in vocational rehabilitation services. CWs grant extensions if criteria such as severe illness or unclear prognosis are met (40). Citizens are required to meet with their assigned CW within the first eight weeks of sick leave. Before the meeting, citizens submit a brief self-report describing the reason for their absence. Transitioning from work to sick leave is not contingent on a specific diagnosis or prior contact with healthcare providers, and formal medical documentation is not initially required. However, CWs may request additional health information from the citizen’s GP or other healthcare providers through formal medical certificates before or after the meeting. It is mandated by law that CWs schedule at least 4 sick leave follow-up meetings over 22 weeks. These meetings primarily focus on citizens’ ability to work and whether any health or social issues should be addressed to promote this goal (41). Furthermore, the meetings aim to promote work participation through work-related and healthcare activities, such as employer dialogue advice, job-seeking activities, and training or treatment (40).

The distribution of tasks and responsibilities among citizens, CWs, and GPs reflects the intersectoral nature of the sick leave system. Citizens are expected to participate in agreed-upon activities to facilitate their return to work and maintain eligibility for benefits. CWs hold a dual role, supporting return to work and determining eligibility for benefits and services. Similarly, GPs have three main responsibilities for people on sick leave: 1) serving as primary care providers offering treatment and counselling, 2) acting as gatekeepers for other health services, e.g., referral to a psychologist or psychiatrist, and 3) documenting citizens’ health status by writing responses to sick leave certificate requests from the citizen’s CW. While GPs can inform decisions regarding entitlement to sick leave benefits through their responses to sick leave certificate requests from CWs, the final decision rests with the CW.

#### 5.2.2 CASE STUDY MUNICIPALITIES

The study was conducted in two municipalities (municipality s) in the Capital Region of Denmark, purposefully selected to assess whether similar patterns would emerge across different contexts, i.e., literal replication logic (42). Municipalities were recruited based on the following criteria: (1) gaining access to a diverse sample of participants for interviews due to a differing sociodemographic profile, (2) enhancing transferability of findings given a varied municipal approach to vocational rehabilitation. Compared with municipality 1, the population served in municipality 2 had, on average, lower socioeconomic resources (e.g., lower average incomes and higher unemployment rates). In turn, the provision of vocational rehabilitation services in municipality 2 relied more on private contractors, such as work placements, resulting in fewer in-house services and a lower degree of interdisciplinarity among municipal personnel compared to municipality 1. The first author approached both municipalities via email, and an in-person meeting with management was scheduled to discuss the study’s purpose, field access, and mutual expectations before the study’s commencement.

### 5.3 DATA GENERATION

We primarily generated data using semi-structured individual interviews with citizens and GPs, as well as focus group interviews with CWs. Interviews were preceded by participant observations of citizens and CWs in the sickness benefits section of the job centre, which served to recruit participants and build rapport, inform the content of interview guides, and contextualise the analysis.

#### 5.3.1 PARTICIPANT OBSERVATIONS AND DOCUMENT ANALYSIS

The first author conducted participant observation for 32 days in the two job centres, observing daily routines, participating in meetings and informal discussions among CWs, and attending CWs’ sick leave meetings with citizens. Additionally, the first author analysed documents, e.g., CWs’ conversation templates, doctors’ sick leave certificates, and judicial documents. Findings were compiled in field notes.

#### 5.3.2 SAMPLING AND RECRUITMENT FOR INTERVIEWS

We recruited citizens and CWs for interviews during the participant observations in the jobcentres. Concurrently, we recruited GPs by outreach via union websites, research network contacts, recommendations from CWs, and “cold calling” GP practices in the two municipalities (43). CWs and GPs were not recruited based on their involvement in care or case management for the interviewed citizens. Consequently, participating GPs were not linked to the interviewed citizens. However, because citizens were recruited via CWs, a few CWs participating in the focus groups were involved in the case management of the interviewed citizens.

Citizens were eligible for interview if 1) they were on sick leave due to common mental disorders, 2) were on long-term sick leave defined as more than three months of sick leave, and 3) were perceived to be at high risk of continued long-term unemployment by their CW. Citizens were recruited via CWs, who identified potential candidates through their case files and, following citizens’ consent, either invited the first author to attend the citizen’s next meeting or shared the citizen’s contact details with the first author. Although there is no universally accepted definition of long-term sick leave, it is commonly defined in the literature as sickness absence lasting 30 days or longer (44,45). We set a threshold of three months’ sick leave to allow CWs to have met citizens on several occasions and to have retrieved relevant health information from the citizens’ GPs. We deemed it important for CWs to make an informed assessment of citizens’ prospects for return to work (the third eligibility criterion). Citizens were strategically sampled for variation in age, sex, sick leave duration, education, and labour market history.

CWs were selected based on variation in age and experience with sick leave cases. Recruitment was coordinated through meetings with the job centre management. CWs were divided into groups according to their work experience to minimise potential hierarchical barriers and promote open discussion.

GPs were selected based on variation in age, sex, experience, and clinic size.

The first author conducted individual interviews with seven citizens, six focus group interviews with 31 CWs, and seven interviews with GPs across the two municipalities (**table 1a-1c**).

Data generation was guided by the principle of information power (46,47), whereby recruitment and analysis proceeded iteratively until we considered the sample to provide sufficient depth, variation, and insight to address the research question. Initially, the study aimed to explore both positive and negative experiences of collaboration among citizens, CWs, and GPs. To this end, we anticipated recruiting approximately 5–10 citizens, 5–10 GPs, and 15–20 CWs. Preliminary analysis indicated that addressing both aspects would exceed the scope of a single article; therefore, the aim was refined to focus on collaborative challenges. Following this narrowing of the analytical focus, recruitment and analysis proceeded iteratively until we judged that the sample provided sufficient information power to address the revised research question. The final number of CWs exceeded our initial expectations because all CWs in the participating sickness benefit departments had been scheduled to attend focus group interviews. As participation had already been organised locally, all six focus groups were conducted. Interviews were audio recorded, and the first author took notes. Student assistants transcribed the interviews verbatim following a transcription guide. The first author subsequently double-checked the accuracy of transcripts by listening to the audio files during coding.

#### 5.3.3 INTERVIEWS WITH CITIZENS, CASE WORKERS, AND GENERAL PRACTITIONERS

Based on participant observations in the job centre, we developed and pilot-tested an interview guide with two GPs having expert knowledge about the collaboration between general practice and municipal job centres and one CW from municipality 2. The pilot revealed the need for tailored interview guides for each actor group and for refining which themes and areas to discuss, leading to the addition of three interview exercises (**Interview guide**).

For CWs, we learned from participant observations that they were accustomed to discussing complex sick leave cases with one another daily. In turn, the individual pilot interview with a CW revealed that differences in language use, understanding of health and work disability, and the societal status of CWs’ social and professional backgrounds, compared with the first author’s medical background, would likely limit the quality of the dialogue in individual interviews. Therefore, we chose to conduct focus group interviews with CWs, as we judged this would likely create a more comfortable atmosphere akin to their daily conversations about complex sick leave cases, while reducing the influence of professional differences between the first author and interview participants when speaking in a group. In addition, the use of focus groups allowed us to include perspectives from more CWs than we would have with individual interviews.

The first author conducted individual semi-structured interviews with citizens and GPs, lasting 92 to 113 minutes and 54 to 90 minutes, respectively. In parallel, the first author conducted focus group interviews with CWs, lasting between 58 and 124 minutes. In both individual and focus group interviews, participants were asked about their experiences with collaboration in vocational rehabilitation for people on long-term sick leave due to common mental disorders. To support reflection, three structured exercises were included in interviews. The first exercise involved reflecting on predefined themes in conceptual pairs, e.g., Trust/Distrust, Order/Chaos, which served as sensitising concepts to guide reflection in interviews about potential tensions in vocational rehabilitation. Themes were derived from the scientific literature and the preparatory phase of fieldwork and pilot interviews. The second exercise concerned comparing real-world collaboration with the ideal definition of vocational rehabilitation. In the third exercise, participants completed a matrix assessing their experience of the involvement (high or low) and contributions (positive/negative) of key actors in vocational rehabilitation (**Interview exercises**).

Interviews with citizens began with open questions about their experience transitioning from work to sick leave, followed by a semi-structured conversation based on the interview guide and exercises. Interviews took place at a location of the citizens’ choice, typically the job centre or the interviewer’s workplace at Frederiksberg Hospital in the Capital Region.

Focus groups with CWs were held at the municipal job centre. The sessions were moderated by the first author, who began by setting ground rules to ensure a safe discussion space. This was followed by group reflections and semi-structured dialogue guided by visible topic cards (**Interview guide**). In both municipality 1 and municipality 2, a co-author was present during the first of three focus groups to observe participants’ interactions and assess the quality of the dialogue and interview approach. Because the focus groups were deemed to function well, only minor adjustments were made to the interview guide and exercises for the subsequent four focus groups, which were conducted without a co-author present.

Interviews with GPs took place in their clinic. They began with open reflections about their role and tasks related to the target group, followed by questions based on the interview guide.

### 5.4 ANALYSIS OF DATA

The analytical approach was rooted in pragmatism, prioritising practical relevance, contextual understanding, and the flexible use of qualitative methods, e.g., making methodological decisions based on what was considered most relevant to practice (48).

Interview transcripts were coded in NVIVO ver15 by the first author using systematic text condensation, an established method for analysing qualitative data (49) rooted in the Scandinavian qualitative research tradition, emphasising open, data-driven exploration of participants’ experiences without applying predefined theory in the initial analytical stages (50). We adapted the method by organising meaning units into subthemes based on the embedded sub-units of the case (e.g., citizens, CWs, and GPs) to explore differences in how each group experienced vocational rehabilitation for long-term sick leave due to common mental disorders. Field notes from observations, document analyses, and interview notes were compiled in a Word document, which was used to contextualise interview findings through iterative comparison, supporting the production of context-sensitive knowledge (48).

To further develop and refine the inductively derived themes, we engaged in an abductive analytical process, iteratively relating our findings to potentially relevant theoretical perspectives (51). Through this process, we chose relational coordination theory as the most suitable framework among the theories considered to support the analysis.

Relational coordination theory posits that collaboration consists of a mutually reinforcing interaction between communication and work relationships among people to facilitate task integration. Further, effective work relationships are characterised by three relational dimensions: a high degree of shared goals, shared knowledge, and mutual trust and respect, which interact with high-quality communication that is frequent, timely, and accurate, with a problem-solving focus (52– 54). To our knowledge, relational coordination theory has been sparsely used in the field of vocational rehabilitation, with only one qualitative study from Denmark using the theory as a lens to interpret inductive findings concerning how GPs, social workers from municipality job centres, and hospital staff experience interprofessional coordination for patients with chronic widespread pain (55). In other fields, the theory of relational coordination has proven suitable for describing contexts characterised by interdependent tasks, uncertainty, and time sensitivity, which we believe is well aligned with the context of vocational rehabilitation (53).

Through an iterative process between theory and data, we used relational coordination theory to interpret and refine emerging themes relating to relational and communication challenges, using the three relational dimensions and the characteristics of high-quality communication as sensitising concepts to examine how stakeholders experience and navigate collaborative challenges (56).

### 5.5 ETHICS

Based on our application for ethical approval of the study, the Regional Committee on Health Research Ethics in Denmark for the Capital Region decided that the study did not require ethical approval (case number F-22051534). Still, the study was conducted in compliance with the Declaration of Helsinki (57). Participant observation of daily routines, meetings, and informal discussions among CWs was negotiated with and approved by management in the municipal sick leave departments. CWs were informed about the purpose of the observations and that they could request an exemption at any time or ask the researcher to leave specific meetings or situations. The researcher did not access any personal or confidential information about citizens, including records or medical certificates. The first author obtained contact information from citizens after they had provided oral consent to their CW to be contacted by the first author. Before interviews, all participants received oral and written information about the study’s aims and methods and how their data would be handled in compliance with the GDPR and the University of Copenhagen’s data storage policy. Participants gave consent by filling in a consent form. Specifically, citizens were informed that choosing whether to participate in interviews would not affect their sick leave process or their care. That information from interviews would not be shared with CWs or other care providers. For CWs, their participation in focus group interviews was supported by management, yet it was explicitly voluntary, allowing for withdrawal or non-attendance without explanation. Likewise, GPs were free to choose whether to participate. Descriptive participant information reported in the study was collected directly from participants with their consent.

## 6 RESULTS

We found that collaborative challenges among citizens, CWs, and GPs were rooted in weak relational ties, characterised by a lack of shared goals, limited shared knowledge, and low levels of mutual trust and respect. Each of these relational challenges was associated with distinct collaborative experiences. Specifically, a lack of shared goals made collaboration feel like a competition for the agenda, limited shared knowledge made collaboration feel laborious, and low mutual trust and respect led participants to perceive one another as adversaries. To navigate these challenges, participants engaged in distinct communication strategies, which we have termed campaigning, clarifying, and self-protective communication. However, rather than resolving the challenges, these communication strategies often reinforced weak relational ties and negative collaborative experiences, contributing to self-sustaining cycles of poor collaboration (**Figure 1**).

**Figure 1.**
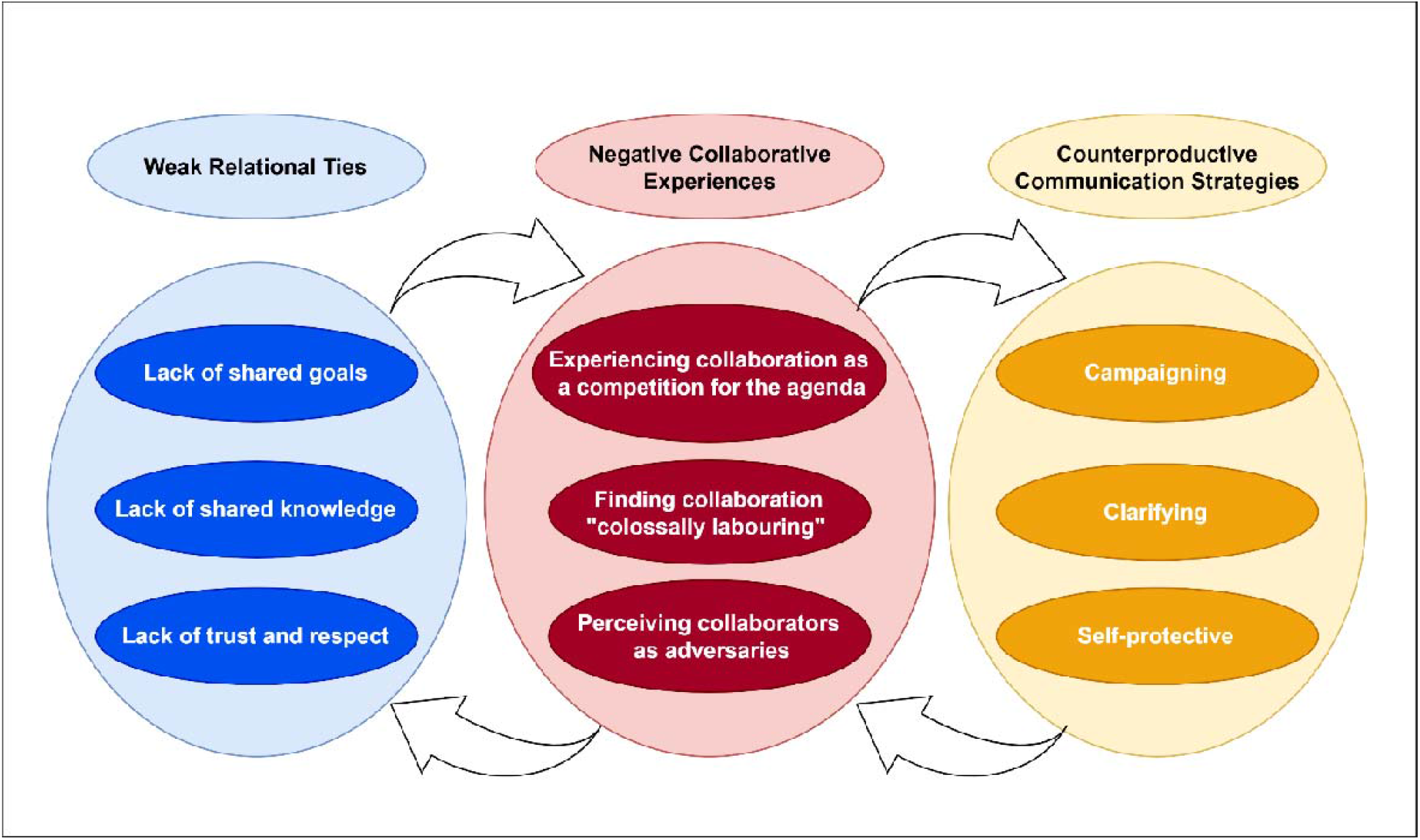
Collaborative Challenges in Vocational Rehabilitation Due to Weak Relational Ties, Negative Collaborative Experiences, and Counterproductive Communication Strategies: An explanatory framework. The findings are presented in three themes corresponding to the three types of weak relational ties shown in Figure 1. Each theme describes: (1) the nature of the relational challenge, (2) the negative collaborative experiences associated with it, and (3) the communication strategies participants used to navigate these experiences.

### 6.1 WITHOUT SHARED GOALS, COLLABORATION WAS EXPERIENCED AS A COMPETITION FOR THE AGENDA

At first glance, it appeared that citizens, CWs, and GPs shared a common goal to increase citizens’ work participation. However, this apparent consensus concealed underlying disagreements about the means and ends of vocational rehabilitation (**table 2**). As a result, the key actors would sometimes pursue divergent functional goals to optimise their agendas at the expense of working towards a shared goal, i.e., subgoal optimisation, resulting in collaborative challenges among them (**table 3**). In general, a conflict emerged between the logic of care, i.e., supporting people according to their needs, and the logic of workfare, i.e., making benefits and access to care services conditional on work or on participation in work-related activities.

#### 6.1.1 CITIZENS PURSUED ACCESS TO CARE AND EXEMPTION FROM WORKFARE

All interviewed citizens expressed a desire to return to work. However, they often found work-related activities, e.g., job placements or sanctions, too demanding to manage due to their health issues. As a result, citizens sometimes experienced activities aiming to promote their return to work as coercive or counterproductive. Instead, citizens primarily sought access to care-related activities, such as illness management support, which they perceived as a prerequisite for participating in return-to-work activities. Thus, citizens perceived the logic of care as superior to the logic of workfare to get back to work, emphasising being seen and being involved as opposed to having to comply with the system’s expectations of participating in various work-readiness activities before being granted access to healthcare services:

…*people who see you and not just a number*… *It gives me space to be me. This is my plan for getting better. What am I feeling? Am I ready to work more? It’s always my need*… *It’s not them dictating something with a stepwise model. You have your life in your hands. You’re the one who decides how it’s going to go. Citizen 57, municipality 2, 1658 days of sick leave*

However, citizens’ logic was at odds with the work-related demands embedded in the Danish sick leave system, known as the work-first principles of active labour market policies. These included having to attend various meetings and other work-related activities to receive sickness benefits despite your health problems. The conflict between citizens’ logic of care and the system’s logic of workfare created a dilemma for citizens regarding the pace of service delivery. On the one hand, citizens wanted peace and the time to heal, i.e., care activities, but on the other hand, they also wanted to receive relevant return-to-work activities that increased their chances of returning to work, exemplified by the following reflection from a sick-listed citizen:

*I like that I get the time to have peace for myself to heal. But sometimes it takes a bit too long before the next thing*… *Sometimes it’s three to four weeks before the next meeting with anyone*… *where I just sit and do nothing. Where I could have healed*… *and got a job again, and be useful*… *Yes, it could be a bit more frequent, but I also think I need that peace. (Citizen 43, municipality 1, 187 days of sick leave)*

#### 6.1.2 CASE WORKERS USED WORKFARE PRINCIPLES TO CARE FOR CITIZENS

The agenda for CWs was to improve citizens’ chances of returning to work by encouraging them, their GP and other stakeholders involved to shift their mindset from focusing on illnesses, limitations, and problems towards seeing possibilities for enhancing citizens’ work ability through internships, training, and other work-related activities, i.e., a workfare logic. This logic was evident in how CWs described their key task as *finding people’s resource tracks, preventing passiveness*, and emphasising concepts such as *work, motivation* and *activeness*, illustrated by the following quote from a young CW:

> … *but then we found that she had some motivation for something, and that was to study Asian culture. So, there is always something you can work with… Everyone has a resource track. I believe it. You must move them from the problem track to the resource track*… *there must be something, right? I believe that. Everyone can do something… everyone has some resources. They just need to be found. (Less experienced CWs, municipality 1)*

Notably, CWs recognised the importance of facilitating access to care-related activities to improve citizens’ health, for instance, by accompanying them to health services or reminding them of scheduled appointments. Still, when CWs supported citizens in their care activities, it was not regarded as an alternative to return-to-work activities. Rather, CWs tried to infuse the principles and key ideas from the logic of workfare, e.g., motivation for work, activeness, focusing on possibilities instead of limitations, into citizens’ care activities. In this sense, for CWs, promoting the logic of workfare was regarded as the best way to care for citizens.

#### 6.1.3 GENERAL PRACTITIONERS PROMOTED CARE TO MAKE SENSE IN WORKFARE

Although GPs shared CW’s emphasis on supporting citizens’ return to work, their approach differed from CWs’. Unlike CW’s logic of workfare, GPs’ agenda was to promote appreciation of the interrelationship between health and work among citizens and CWs by emphasising the work benefits of health services and vice versa, the health benefits of work services. Therefore, GPs would often argue for the value of return-to-work activities if these were perceived to be designed and delivered in a way that took account of people’s health problems and their whole life circumstances. One GP explained her approach as such:

*Citizens need some “counterbalancing” to understand that staying at home, waiting until you’re well enough doesn’t make you healthy*…*we help them by saying that they need to be a little more exploratory*… *returning to work is something you can do gently and gradually… start by having a cup of coffee with your colleagues. That’s how we dedramatize and demystify getting back to work. That’s good medicine… (GP33, municipality 1, 23 years of experience)*

Thus, GPs emphasised the interrelationship between health and work by framing health services as supporting work ability and by advocating for work-related measures as beneficial for health.

#### 6.1.4 CAMPAIGNING FOR ONE’S OWN LOGIC IN THE ABSENCE OF SHARED GOALS

The lack of a shared logic regarding the means and ends of vocational rehabilitation led the key actors to engage in campaigning, a communication strategy focused on persuading others of one’s own logic:

*… some citizens say things like: “I just need peace and quiet”. Then we need to have that conversation about it’s not passive support*… *and they say: “my doctor has said that I just need to take it easy”. But you can’t do that. That’s not part of Sick leave legislation. (Less experienced CWs municipality 1)*

Interestingly, the key actors described their own communication in terms that aligned with the characteristics of high-quality communication in relational coordination theory, i.e., frequent, timely, accurate, and problem-solving communication. However, comparing the key actors’ campaign examples revealed that high-quality communication from one perspective was often perceived as low-quality from others’ perspectives. For example, citizens described their campaigning efforts to convince others about access to care and exemption from workfare in terms aligned with high-quality communication. However, these citizen campaigning efforts were frustrating for CWs, who perceived them as low-quality communication. Similarly, what CWs perceived as poor or unhelpful communication in sick leave certificate responses from GPs was conversely considered good communication quality by many GPs.

### 6.2 WITHOUT SHARED KNOWLEDGE, COLLABORATION WAS EXPERIENCED AS COLOSSALLY LABOURING

Collectively, citizens, CWs, and GPs reported a lack of shared knowledge about each other’s expertise, working conditions, and role (**table 3**). The lack of shared knowledge among the key actors caused misalignment in mutual expectations about the responsibilities, rights, and obligations of various stakeholders within the sick leave system. In turn, citizens highlighted a general lack of understanding among professionals regarding their lived experiences as sick leave recipients. This absence of shared knowledge caused distinct types of collaborative challenges for the key actors: 1) Citizens felt obliged to bridge knowledge gaps between professional, 2) CWs repeatedly had to explain the rules and regulations about sick leave while struggling to retrieve relevant information for the sick leave case, and 3) GPs set clear professional boundaries due to what they perceived were unrealistic expectations from them by citizens and CWs.

#### 6.2.1 CITIZENS FELT BURDENED BY TRYING TO BRIDGE KNOWLEDGE GAPS

Citizens were often unsure who to contact about specific issues or how professionals coordinated their cases, expressing a need for clearer role definitions among the many actors involved:

*… What is your role for me? You shouldn’t play half psychologist, half doctor or half caseworker. That’s who you are. And that’s your role, rather than everyone overlapping. (Citizen 27, municipality 1, 206 days of sick leave)*

Further, citizens often reported a mismatch between the high cognitive demands of navigating the sick leave system and their reduced cognitive capacity due to stress and mental illness. These experiences were exacerbated by the poor continuity in case workers responsible for their sick leave case, forcing citizens to explain and repeat their sick leave story multiple times without knowing how their story would be received, i.e., whether they would feel understood and helped:

*It was just that during the same case, I could have a new caseworker every month. And then I had to explain things again. And then it was almost like winning the lottery if someone was sympathetic to the story. And it wasn’t because I wanted to leech off society, I just wanted help. (Citizen 43, municipality 1, 187 days of sick leave)*

#### 6.2.2 CASE WORKERS COMPENSATED FOR OTHER PEOPLE’S SCARCE KNOWLEDGE

Although explaining sick leave regulations was formally part of CWs’ role, they found it time-consuming and, at times, unfair when it extended beyond what they considered reasonable. CWs described a mismatch between citizens’ and GPs’ expectations regarding providing extensive guidance on rules and procedures, while simultaneously struggling to obtain relevant information about citizens’ work capacity from both citizens and GPs. CWs attributed this issue to differing degrees of knowledge about the sick leave system, which could lead to resistance from citizens and GPs in the exchange of sick leave information:

*We have legislation that we must follow. Sometimes we feel that people try to sneak around it. There’s nothing better than talking about yourself and your illness. But our conversations are not meant to concern illness. It’s about how to get back to work*… *(Experienced CW, municipality 2)*

#### 6.2.3 GENERAL PRACTITIONERS SET BOUNDARIES TO MANAGE EXPECTATIONS

GPs often felt that citizens and CWs had unrealistic expectations of them, e.g., CWs requesting definitive answers to complex conditions like stress or mild depression. In such cases, GPs struggled to convey what they felt were the necessary nuances of sick leave cases, being concerned that their assessment would be interpreted as definitive answers:

*Predicting when people’s treatment will be optimised, if all treatment options have been tried, and when people can return to full work ability*… *Whatever that means… It’s like we’re supposed to be the oracle in Delphi, right? The idea that case workers think they can use us for that*… *It’s a symbol of a general juridification of society… thinking that everything can be answered just by asking*… *I don’t mind being asked, but don’t use it as a fact. Use it as an opinion, as an information piece. Then they must judge what it means for people’s ability to work. (GP 49, municipality 2, 5 years of experience)*

A similar concern for GPs was when citizens expected validation of their perceived care needs and exemption from workfare, which did not align with GPs’ clinical judgment, pushing GPs towards setting clear boundaries around their professional role:

*My role is to be a doctor*… *I’m not the judicial power. Sometimes, citizens tell me that the job centre has advised them to write that they need an early retirement pension. Then I tell them: “No, I’m not going to do that. I must examine you and explain what’s wrong with you, as well as what we can do to make you as well as possible. It’s not my job to find jobs for them or to judge them. (GP 42, municipality 2, 21 years of experience)*

#### 6.2.4 CLARIFYING COMMUNICATION PARTIALLY COMPENSATED FOR THE LACK OF SHARED KNOWLEDGE

The key actors tried to navigate the low degree of shared knowledge via *clarifying* communication strategies. These strategies aimed to reduce ambiguity and confusion within the collaboration by explaining one’s situation, perspectives, self-perceived role, and expectations regarding others’ roles and tasks. However, *clarifying* communication strategies only partially compensated for the low degree of shared knowledge among the key actors. Further, the price of these strategies was extra work and a collective experience of the collaboration as *“colossally labouring”*, as one CW exclaimed during an interview.

Citizens’ primary clarification strategy was to act as *messengers*, relaying their sick leave stories to the various stakeholders involved in their case. Paradoxically, this “citizen work” reduced citizens’ energy to engage in rehabilitative activities such as socialising, exercise, or hobbies:

> … *I focus on all this stuff with the job centre, doctors and what’s going to happen, and work, and I put my energy into that. But when I get home, I don’t have the energy for doing anything else*… *there’s simply no room for it. I feel sad to say it out loud. It’s just hard to be honest about it. It takes me a great deal of mental effort*… *I’m just completely exhausted, right? (Citizen 27, municipality 1, 206 days of sick leave)*

In turn, CWs adopted the role of “bureaucracy guides”, feeling responsible for repeatedly explaining sick leave rules to citizens and GPs, having to guide them about services and regulations to a degree that they felt was outside their formal responsibilities:

*… and people say things like: ‘Well, I’m ill’. Yes - but what does it take for you to get back to work? We don’t focus on how many band aids they’re wearing. That’s the first thing they need to understand. (Experienced CW, municipality 2)*

For GPs, clarifying communication involved a more narrowly focused and standardised approach towards people on sick leave than they typically took when dealing with patients in primary care. Here, their first step was to explain to citizens or CWs what they considered were the medical versus the non-medical problems related to sick leave, which some GPs described as “closing the somatic door for the patient”:

> … *the reason I take all these blood tests is that they often fear something serious is wrong with them. At the follow-up appointment, we can say: See, you’re not seriously ill, you’re not dying*… *and then we can get back to the psychological part. (GP 42, municipality 2, 21 years of experience)*

Second, GPs zoomed in on managing medical issues:

> *My role is to clarify what’s going on. Optimise as quickly as possible*… *And if there’s something wrong, we need to find out what it is. And is it something we can do better? (GP 33, municipality 2, 23 years of experience)*

Third, GPs signposted citizens to other service providers outside of the health system to support the fact that these issues were also taken care of:

> *I think we help them by explaining that this isn’t a medical issue. If it’s stress, or something psychological, or something related to their employer. An imbalance in their lives it’s about separating the wheat from the chaff to find out what’s medical in this and take care of that. (GP 49, municipality 2, 5 years of experience)*

This three-step strategy helped GPs maintain professional boundaries and avoid being misunderstood.

### 6.3 WITHOUT MUTUAL TRUST AND RESPECT, COLLABORATORS WERE PERCEIVED AS ADVERSARIES

Low levels of trust and respect among citizens, CWs, and GPs emerged as a major driver of collaborative challenges. The main consequences of mistrust and disrespect were that the key actors began to perceive their collaborators as obstacles or adversaries, because they felt ignored, dismissed, misunderstood, or actively opposed in their needs, views, contributions, or recommendations for the sick leave case (**table 3**).

#### 6.3.1 CITIZENS EXPERIENCED A LACK OF SICK LEAVE VALIDATION FROM PROFESSIONALS

Sharing their stories about sick leave, many citizens explained how they sometimes felt misunderstood, blamed or disrespected by professionals who did not validate their experiences, opinions, or needs; for example, when professionals were unwilling to support their requests for access to care or pressured them into work-related activities, which they felt unable to perform. This, in turn, created a sense of mistrust of the professionals among the citizens. As a result, they sought trustworthy professionals who were good at listening to their story, offering support, taking the required time, and believing their account, e.g.:

*They care for me and support me. It’s not that I need them to echo my words, but they agree with my assumptions*… *It just feels safe*… *They confirm that what I’m feeling, thinking and experiencing is okay. (Citizen 31, municipality 2, 195 days of sick leave)*

Citizens described this as a human approach that affirmed the legitimacy of their experiences, which they contrasted with a system-driven approach characterised by system priorities taking precedence over their needs, e.g., reducing the duration of sick leave as much as possible rather than allowing time for relevant treatment and improvement in functioning.

#### 6.3.2 CASE WORKERS FELT MISTRUSTED, DISRESPECTED AND UNDERVALUED

CWs experienced negative media narratives about job centres had a significant impact on how others viewed them. Some CWs even hesitated to discuss their work with friends or family due to fear of criticism. Hence, CWs felt that citizens and GPs often mistrusted their intentions due to their role as job centre representatives:

*… the media is just out to get us, right? Whether it’s social media*… *on TV*… *or wherever it is*… *oh my God, we’re so reviled. Do you think there’s anyone in Denmark who doesn’t know what the Job Centre’s Victims are (a patient organisation)? Where do all the good stories go, I wonder, because God knows there are many of those also. (Less experienced CWs, municipality 1)*

The challenge related to gaining trust from GPs and citizens was especially pronounced for less experienced CWs, who struggled to balance their dual role of providing support and guidance while assessing and deciding on eligibility for sickness benefits and other services:

*It can be very difficult to wear both hats, offering support, guidance and counselling, but also wearing the authority hat and advising about consequences and sanctions. It can make it difficult to build trust. (Less experienced CWs, municipality 1)*

However, experienced CWs were also affected by the low degree of trust and respect from citizens and GPs, having to earn trust from a very low starting point in their initial encounters:

> … *most people meet us with prejudices. Especially if it’s their first time here or if they’ve had a bad experience before. (Experienced CW, municipality 1)*

Further, many CWs expressed that they felt at the bottom of a professional hierarchy, with their judgments frequently dismissed in favour of those from health professionals, even on matters within the caseworkers’ expertise:

*… if your GP says you should never work again, they can’t hear anything else. If we say something else, then there is no respect for that*… *Or if it’s a doctor at the hospital or something, they’ll always rank higher than us. It’s like it’s the law if the doctor has said so, but here at the job centre*… *No, they won’t listen to us. (Experienced CWs, municipality 1)*

While experiencing mistrust and disrespect from others, CWs themselves tended to lose trust and respect in GPs who were perceived as uncritical about how they handled sick leave certificates. In such cases, CWs questioned whether GPs genuinely assessed cases or merely echoed citizens’ accounts, which was described as *“holding the microphone for the patient” (Experienced CWs, municipality 2)*. CWs’ trust and respect for citizens and GPs were further strained when GPs or citizens appeared to disregard their contributions, or when citizens and GPs focused solely on illness and limitations rather than possibilities for a return to work.

#### 6.3.3 GENERAL PRACTITIONERS’ FEAR OF BEING MISUSED FOR QUESTIONABLE CLAIMS

Unlike CWs and citizens, GPs rarely felt that others mistrusted them. However, they sometimes felt disrespected when their medical expertise was misunderstood or misused. First, GPs mentioned encounters with citizens who had appeared to exaggerate symptoms or attempted to influence the content of sick leave certificates to avoid work-related activities, leaving GPs feeling misused in legitimising questionable claims. Second, GPs sometimes felt that CWs misused their assessments to justify untimely or unjustified closures of sick leave cases, or that their care-related recommendations were ignored. In more extreme cases, GPs would lose respect for CWs when they received sick leave certificate requests that seemed automated, uncritical or unethical. For example, GPs had little respect for CWs who sent a template of 10 standard questions without any tailoring or concern for the individual citizen. Conversely, GPs did not appreciate it when CWs returned their certificates, asking for more professional assessments, implying that the GP had not been sufficiently thorough or was somehow biased in their assessment. In such cases, many GPs felt compelled *“to put their foot down”* or “*to stand up for what was right for their patient”*:

*I’ve had a patient who was on sick leave due to stress and anxiety, and then she got breast cancer… And you could tell that the caseworker was just sick and tired of the process and questioned whether my patient was ill and asked when we could send her back to work… I had to write back… she has cancer. Back the f*** off. She had family, friends, and two small children at home. Everything was disintegrating*… *I had to write it so the social worker on the other end understood that it’s not one-size-fits-all. (GP 58, municipality 2, 8 years of experience)*

#### 6.3.4 SELF-PROTECTIVE COMMUNICATION STRATEGIES UNDERMINED TRUST AND RESPECT

When trust and respect among citizens, CWs, and GPs were lacking, they engaged in various self-protective communication strategies to shield themselves against perceived threats from their collaborators, such as criticism. However, these *self-protective* communication strategies unintentionally weakened relational ties among the key actors, undermining effective coordination concerning the sick leave case.

Citizens’ primary self-protective strategy was to *camouflage*, i.e., carefully curating what to say and to whom. This strategy served multiple purposes, including a) to safeguard their relationships with their CW, GP and other stakeholders involved in their case, b) to align their sick leave story with demands for receiving sick leave benefits, c) to promote access to care-related activities, e.g., speaking to a psychologist, and d) to be exempted from work-related activities that were perceived as unhelpful, e.g., telling their sick leave story in a way that showed their inability to take part in a work placement. Unfortunately, when professionals perceived that citizens might be withholding or masking important information, this undermined the relationship and reduced trust and respect. Importantly, citizens felt compelled to navigate the system this way, by putting up a façade or performing a role, which was an approach they found uncomfortable:

*… well, when I said I feel okay, I mean I do have periods where I feel good sometimes, but then the next day, I go completely downhill*… *When I told my caseworker, they wrote it down and thought they could send me to all kinds of work-related activities. So, I’m very careful about what I say now… And it’s uncomfortable that you must sit there and keep quiet, not expressing your honest feelings. I want to express how I feel, but I’m afraid that what I say will be used against me. (Citizen 75, municipality 2, above two years of sick leave)*

As opposed to citizens’ defensive camouflaging communication strategy, CWs tended to adopt an authoritative communication style if they felt a lack of trust and respect from their collaborators. Here, CWs would emphasise their legal mandate to retrieve information, or explicitly list what was expected from a GP in a sick leave certificate, or the eligibility criteria for receiving sickness benefits for citizens. Further, CWs would try to reframe conversations with citizens or GPs from talking about citizens’ health issues towards citizens’ potential for returning to work, including challenging vague medical documentation from GPs by requesting “more objective assessments”:

*It’s not uncommon for us to send a medical certificate back to the GP saying: “Fine, now we can see that you’ve written what the citizen says, but what do you think? What is your assessment?” It is a <u>medical</u> assessment*… *can you give us one of those? Instead of what the citizen thinks. (Experienced CW, municipality 2)*

GPs’ self-protective communication strategies ranged from avoidance and indirect communication to more assertive approaches to protect their professional integrity and/or their patients, such as deflecting responsibility or setting clear boundaries. Regardless of form, these strategies inadvertently undermined relationships with CWs and citizens.

Some GPs described the use of vague or *“elastic language*” in sick leave certificates to avoid legitimising questionable practices while advocating for their patients’ needs:

…*We often write a bit in “elastic terms” Because we’re a bit afraid that this must be understood as being closer to an opinion than a fact because if there’s a field in the medical certificate like how many months and weeks until expected full working capacity, then you can’t answer, ‘probably around… and must be reassessed continuously’. (GP 49, municipality 2, 5 years of experience)*

In addition to this *“elastic-type language*”, these GPs would tend to adopt what they called an “*ostrich approach*,” aligning their recommendations with their patients’ preferences to preserve the therapeutic relationship with their patient, e.g., extending the sick leave period solely based on how citizens experienced their limitations:

> … *Often you sound like a bully just mentioning return to work - So sometimes I stick my head in the sand*… *I’m a bit of an ostrich about it. Because people don’t have to lose their GP in the process*… *(GP 33, municipality 1, 23 years of experience)*

Conversely, other GPs described more assertive communication in sick leave cases where they had felt obliged to *“put their foot down”* or *“stand up for what was right*”, even if it meant stepping into CWs’ professional domain, e.g., by recommending early retirement pension or provision of social services for citizens. Consistent with this approach, these GPs often left the decision about sick leave to the patient to safeguard their professional role as medical experts, i.e., delegating the decision about sick leave to the citizen, a strategy they referred to as “*passing the buck*”:

*We no longer put people on sick leave. I certainly don’t. I tell citizens that going on sick leave is their decision – not mine. (GP 49, municipality 2, 5 years of experience)*

## 7 DISCUSSION

### SUMMARY OF KEY FINDINGS

This study explored how citizens on long-term sick leave due to common mental disorders, CWs, and GPs experience and navigate collaborative challenges in vocational rehabilitation. Using relational coordination theory as an explanatory framework, our findings suggest that collaborative challenges can be understood as self-reinforcing cycles between weak relational ties, negative collaborative experiences, and counterproductive communication strategies. While previous research has identified barriers such as conflicting goals, poor mutual understanding, communication difficulties, and limited trust among stakeholders in vocational rehabilitation (24–37,58–60), our findings extend this literature by illustrating how these barriers interact and are inadvertently sustained through stakeholders’ attempts to manage them in practice.

Specifically, divergent views on the means and ends of vocational rehabilitation contributed to experiences of competing for the agenda, which were reinforced by campaigning communication strategies aimed at persuading others of one’s own perspective. Limited shared knowledge about roles, responsibilities, and expectations made collaboration feel laborious and contributed to the need to clarify communication strategies, which often intensified rather than resolved misunderstandings among the key actors. Finally, low levels of mutual trust and respect gave rise to experiences of being ignored, dismissed, misunderstood, or opposed, prompting self-protective communication strategies that further widened the relational divide between stakeholders.

To our knowledge, no previous studies in vocational rehabilitation have described these collaborative challenges as interconnected and self-perpetuating processes. Further, although relational coordination theory has been widely applied in other fields, a recent review identified only one prior study that applied the theory in vocational rehabilitation (35)(58). By applying relational coordination theory to interpret stakeholders’ experiences, the present study offers an explanatory framework for understanding not only which collaborative challenges occur but also how they are linked to the quality of relational ties, how citizens, CWs, and GPs experience them, how they are navigated via communication strategies, and how the interrelationships between these can make them sustained over time.

#### 7.1.1 ARE SHARED GOALS FOR VOCATIONAL REHABILITATION ATTAINABLE?

Arguably, stakeholders in vocational rehabilitation share many common views, and previous studies have outlined what can be expected from their different roles (28,60). Still, we found that stakeholders’ differing perspectives on the means and ends of vocational rehabilitation can hinder successful collaboration, corroborating findings from previous studies (24,55,60). For example, we found that citizens preferred access to care and exemption from workfare to improve their health and functioning, making a return to work a secondary goal. Similar findings from the Danish sick leave system indicate that citizens do not view workfare measures, e.g., work-first initiatives or sanctions, as helpful for rehabilitation or return to work (61,62).

In turn, we found that a lack of shared goals among stakeholders leads to campaigning strategies aimed at persuading others of one’s view, i.e., competing for the agenda. This undermines agreement on shared goals and sustains subgoal optimisation instead of collective coordination. These findings have important implications for common recommendations aimed at increasing stakeholder involvement in the design and implementation of vocational rehabilitation services. Such recommendations are often based on the assumption that greater involvement promotes shared goals, alignment of expectations, and successful implementation (24,27,63–66). Our findings suggest that this assumption may be overly simplistic. Even when stakeholders are actively involved, collaboration may remain challenging if fundamental disagreements persist regarding the purpose of vocational rehabilitation and the relative importance of care, work participation, and benefit administration. Our findings resonate strongly with Franche et al.’s concept of paradigmatic dissonance (67), which suggests that tensions between stakeholders arise not simply from poor coordination but from fundamentally different professional mandates, responsibilities, and values. From this perspective, complete agreement on the means and ends of vocational rehabilitation may be neither realistic nor necessary. Rather, improving collaboration may depend on creating conditions that enable stakeholders to recognise, tolerate, and work constructively across these differences. The strategies proposed by Franche et al. include clarifying roles, improving communication, and fostering mutual understanding (67), which align closely with our findings and may represent practical approaches to reducing the negative consequences of competing agendas in vocational rehabilitation.

#### 7.1.2 DO RETURN TO WORK COORDINATORS HOLD THE KEY TO SHARED KNOWLEDGE?

Recent studies from Sweden and Denmark similarly highlight both the importance and the challenges of creating a more informative collaborative environment with clear roles, responsibilities, and expectations among vocational rehabilitation stakeholders (24,60). Our findings extend this literature by illustrating how citizens, CWs, and GPs experience limited shared knowledge as a burdensome process of repeatedly having to explain themselves, correct misunderstandings, and navigate uncertainty about one another’s roles and responsibilities. Consistent with previous research, this often positions citizens as intermediaries responsible for transferring information between professionals and organisations (68,69).

One approach to addressing such challenges is to use return-to-work coordinators, professionals dedicated to coordinating and communicating among stakeholders throughout the rehabilitation process (70,71). Studies suggest that these schemes can improve communication, reduce fragmentation, and help balance power relations between stakeholders, although their effects on work participation remain mixed (24,60). In comparison to our findings, return-to-work coordinators may be particularly valuable because they function as boundary spanners, i.e., individuals who bridge professional, organisational, and knowledge boundaries between actors who otherwise struggle to establish a shared understanding (67,70,71). By facilitating information exchange, translating perspectives, and clarifying roles and expectations, boundary-spanning actors may help reduce the collaborative burden associated with limited shared knowledge. However, previous studies also suggest that such roles can challenge existing professional identities and impose additional collaborative demands on already-strained professionals, which may complicate their implementation and effectiveness (24,60). In turn, consensus on best practices for return to work coordinators is currently still lacking (60,72,73)

#### 7.1.3 SHOULD THE SICKNESS CERTIFICATION PROCESS BE REVISED TO FOSTER MUTUAL TRUST AND RESPECT?

Previous studies have highlighted the importance of establishing a supportive collaborative environment in vocational rehabilitation, characterised by mutual trust and respect, open communication, and recognition of stakeholders’ roles, expertise, and constraints (60). Our findings support this recommendation by illustrating how low levels of trust and respect can contribute to experiences of being ignored, dismissed, misunderstood, or opposed, which in turn can lead stakeholders to engage in communication strategies that further widen the relational divide.

The challenges experienced by GPs in navigating competing expectations from citizens and CWs and dilemmas between the interests of the patient, society and their professional integrity resonate with previous studies showing that sickness certification is a complex and value-laden task requiring professionals to balance their responsibilities towards patients, employers, welfare authorities, and society as a whole (74–76).

Similarly, our findings suggest that CWs’ opportunities to establish trusting and respectful relationships are constrained by current organisational and policy conditions, which emphasise work ability and return-to-work outcomes over broader health-related concerns (77,78). Although these policies might support citizens’ labour market participation, they can also restrict CWs’ opportunities to exercise professional discretion and tailor support to individual circumstances. Further, as noted in previous studies, many CWs in our study described feeling mistrusted, disrespected, and positioned at the bottom of a professional hierarchy (79). Efforts to strengthen collaboration may therefore require not only improving communication between stakeholders but also ensuring that CWs have the organisational support, legitimacy, and professional autonomy needed to perform their coordinating role effectively.

Finally, citizens’ opportunities for developing trust and respect towards professionals may likewise be shaped by how vocational rehabilitation services are organised. Previous studies have shown that people on sick leave due to common mental disorders value being listened to, involved in decision-making, treated as credible informants about their own situation, and supported through often complex and fragmented rehabilitation processes (24,25,29,33,34). Consistent with these findings, many citizens in our study described experiences of not feeling heard, understood, or sufficiently acknowledged by professionals. Efforts to strengthen trust and respect may therefore require not only better communication between stakeholders, but also more person-centred and coordinated rehabilitation processes that reduce uncertainty, recognise citizens’ vulnerabilities, and enable meaningful involvement in decisions affecting their rehabilitation (31,36).

### 7.2 STRENGTHS AND LIMITATIONS

Our analytical ambition to explore how citizens on long-term sick leave due to common mental disorders and professionals experience and navigate collaborative challenges in vocational rehabilitation comes at the expense of describing what works well in the Danish sick leave system. Second, limiting eligible participants in this study to citizens, CWs, and GPs excludes perspectives from other stakeholders, including citizens’ relatives, employers, other health and social care personnel, and third-sector organisations, e.g., volunteers. Third, our study does not account for the broader contextual conditions influencing stakeholders’ opportunities for collaboration. While these methodological choices limit the breadth of our findings, they arguably deepen understanding of the dynamics of collaborative challenges on the interpersonal level among key stakeholders directly involved in managing sickness benefits and return-to-work processes in the Danish sick leave system.

Other methodological considerations are outlined below using the four pillars of trustworthiness in qualitative research: credibility, transferability, dependability, and confirmability, as a structure (80).

Credibility, i.e., how well the findings reflect participants’ actual experiences (80), was strengthened by collecting data over an extended period using multiple sources. In turn, as an interviewer, the first author established in-depth access to the field and a strong rapport with participants in both municipalities (42). We attribute this to the first author’s medical background, practical experience as a medical consultant, and prior professional relationships with some of the participants from municipality 2. However, these same characteristics could also challenge the credibility of findings if not addressed reflexively. To avoid conceptual blindness and promote ongoing reflexivity about his role and its implications for the findings, the first author noted his preconceptions before data generation, kept reflective notes in a research diary throughout the study, and discussed these regularly with a multiprofessional research group and clinical colleagues from different professional backgrounds. Further, it was our impression that the sample provided sufficient information power to address the research question for the following reasons: 1) the study’s aim was narrowed down to a focus on collaborative challenges among the three key stakeholder groups in the Danish sick leave system, 2) participants were strategically sampled to ensure variation within each group (table 1a-1c), 3) interview quality was supported by guides informed by prior observations, structured exercises, reflexive journalling, and iterative analysis during data generation, and 4) relational coordination theory provided an analytical framework for interpreting the data following inductive analysis.

Transferability of findings, i.e., the extent to which findings can be applied to other contexts or groups (80), was promoted by purposive sampling of two municipalities with differing sociodemographic and organisational practices (i.e., a literal replication logic (45)) and by high-variation sampling of citizens, CWs, and GPs. Although the study was conducted within the Danish sick leave system, several of the collaborative challenges identified, including competing stakeholder agendas, limited shared knowledge, lack of trust, communication difficulties, and citizens acting as intermediaries between services, have been reported across a wide range of vocational rehabilitation settings internationally (25,27,28,35,37). While the organisational details of vocational rehabilitation systems vary between countries (5,38), the involvement of citizens, healthcare professionals, social, and employment services appears to create similar coordination challenges across settings. This suggests that the explanatory framework developed in the present study may have relevance beyond the Danish context.

Dependability, i.e., consistency and reliability of the research process over time (80), was supported by systematic and transparent reporting of methods using known reporting guidelines (81,82) and having a detailed audit trail of analytic decisions and coding revisions in a reflexive diary kept by the first author.

Confirmability, i.e., the degree to which findings are shaped by participants and not the researcher’s preconceptions (80), was enhanced through by 1) iterative coding and conduct of interviews, permitting revisions to interviews guides and iterative comparison between preliminary themes, field notes and interview transcripts, 2) reflexive journaling by the first author to remain aware of his pre-conceptions about the field, 3) peer debriefing, e.g., presenting and discussing emerging findings with various professionals in the field, e.g., case workers, psychologists, and doctors, and 4) ongoing discussion of emerging findings in the group of authors, facilitating a transparent progression from data to themes, and 5) postponing theoretical perspectives on the data at a late stage of the analysis.

### 7.3 FUTURE RESEARCH

To our knowledge, this is only the second study to apply relational coordination theory in vocational rehabilitation. By using the theory as an explanatory framework for understanding collaborative challenges, the study responds to recent calls for greater use of theory and well-defined concepts from the field of interorganisational collaboration in vocational rehabilitation research (35). Future studies could explore the transferability of the explanatory framework developed here across different vocational rehabilitation systems and institutional contexts.

A second priority concerns the broader organisational and societal conditions that shape opportunities for effective relational coordination. A recent systematic review has demonstrated that relational coordination is influenced by multiple contextual factors operating at organisational and structural levels (53). Future research could investigate how these contextual factors influence the development of shared goals, shared knowledge, mutual trust and communication among stakeholders in vocational rehabilitation.

Finally, our findings suggest that collaborative challenges arise in part from differences among stakeholders’ professional roles, mandates, and cultures. Future studies could examine how these factors shape stakeholders’ understanding of vocational rehabilitation and contribute to increased tolerance of paradigmatic dissonance among stakeholders in vocational rehabilitation. Such knowledge may help inform future interventions, professional training, and service design aimed at strengthening collaboration in vocational rehabilitation.

### 7.4 CONCLUSION

This study explored how citizens on long-term sick leave due to common mental disorders, CWs, and GPs experience and navigate collaborative challenges in vocational rehabilitation. Using relational coordination theory as an explanatory framework, the findings suggest that collaborative challenges can be understood as self-reinforcing cycles between weak relational ties, negative collaborative experiences, and counterproductive communication strategies.

Specifically, a lack of shared goals, shared knowledge, and mutual trust and respect contributed to experiences of competing for the agenda, finding collaboration laborious, and perceiving collaborators as adversaries. Rather than resolving these challenges, stakeholders’ attempts to navigate them through campaigning, clarification, and self-protective communication often reinforced the underlying relational problems and sustained them over time.

These findings suggest that efforts to improve vocational rehabilitation should focus not only on individual barriers to collaboration but also on strengthening relational coordination among stakeholders. This may require organisational conditions that support shared goals, shared knowledge, mutual trust and respect, and high-quality communication across professional and organisational boundaries. The explanatory framework developed in this study offers a novel way of understanding how collaborative challenges emerge and persist in vocational rehabilitation and may inform future research and interventions aimed at improving collaboration.

## Supporting information

Implications for rehabilitation

## Data Availability

All data produced in the present work are contained in the manuscript.

## 8 ACKNOWLEDGEMENTS

The authors would like to thank the participating caseworkers, general practitioners, people on sick leave, and the management teams at the municipal job centres for generously sharing their time and experiences as part of this study. Special thanks to Sofie Fulton for constructive feedback on the preprint before publication.

## Use of Generative AI

During the preparation of this manuscript, the authors utilised OpenAI’s ChatGPT to support language editing, enhance clarity, and improve readability. The authors reviewed all content and take full responsibility for the manuscript.

## 11 APPENDICES

## 11.1 INTERVIEW GUIDES

### 11.1.1 INTERVIEW GUIDE FOR PEOPLE ON SICK LEAVE

| Subject | Question |
| --- | --- |
| Transitioning from work to sick leave | In your own words, what were the main reasons for transitioning to sick leave? What happened then? Was there anything that could have prevented it? Can you talk me through it? |
| Meeting the job centre | How was it for you to get in touch with the job centre?<br>How has it been since then for you to be in the sickness allowance system?<br>Has it changed over time? |
| Self-understanding in relation to transition | Has becoming a sickness allowance recipient affected your self-understanding, i.e. how you see yourself? If so, how?<br>People on sick leave due to common mental disorders have a higher-than-average risk of long-term sick leave, commonly referred to as being “vulnerable”. Do you see yourself as a vulnerable person or as an ordinary person put in a vulnerable situation? |
| Thoughts | What thoughts has been triggered in you from being in the sickness benefit system? Has anything surprised you? Are there things you wish you had known earlier? |
| Emotions | What feelings has been triggered in you from being in the sickness benefit system? |
| Bodily sensations | Have you noticed any bodily sensations during your time as a sickness allowance recipient? Is the experience of being on sick leave connected to the body? If so, how? |
| Coping/Behavior | What do you do as a sickness benefit recipient? What kind of behavior does this require on your part? What skills do you need to have? Do you feel you have the required skill set and resources? |
| Opinion | Is there anything you would like to highlight as particularly good or bad about your sick leave process? |
| Time | How do you experience time? When is there enough time for recovery, when is there too little time? Too much time? What about waiting time? |
| System | Do you feel that the system is transparent and understandable? |
| Security/Trust | Are you comfortable with the people you have around you? Do you trust them to help you? Conversely, are there people whom you may find it difficult to trust? |
| Exercise 1 | Talk about potential themes describing the experiences related to the sick leave process |
| Exercise 2 | Discuss the ideal of vocational rehabilitation |
| Exercise 3 | Do you have people around you who support you in your sickness allowance process?<br>What role do they play and who helps you with what?<br>Do you have any people around you who you feel oppose you? |

### 11.1.2 INTERVIEW GUIDE FOR GENERAL PRACTITIONERS

| Theme | Subject | Question |
| --- | --- | --- |
| The target group | Size of the problem | How much time and effort is required to handle people on sick leave due to common mental disorders in your practice? Has this changed over time? When and why do they come to you? |
|  | Long term unemployment | Why do you think people on sick leave due to common mental disorders tend to become long-term unemployed? From your experiences, are there certain predictive factors for long-term unemployment? If yes, which? |
|  | Vulnerability | People on sick leave due to common mental disorders have a higher-than-average risk of long-term sick leave, commonly referred to as being “vulnerable”. Do you think vulnerability is something “you are”, i.e., concerns personal characteristics, or is something “you become” in certain situations or both? What would you say characterizes those “that are” vulnerable? What is the opposite? How do we promote that instead? |
| Experience with the target group | Sick leave work in general practice | Which tasks are associated with sickness benefit recipients with mental health problems in your practice? What challenges do you face here? |
|  | Role | How do you see your role in relation to the target group?<br>What are your core tasks? |
|  | Approach | Do you do anything special when you work with particularly vulnerable sickness benefit recipients with mental illness? What do you focus on? |
|  | Context | Do you feel that you have the right conditions to help the target group as much as possible? What things promote or hinder you from making a positive difference? |
|  | Knowledge and skills | Do you have sufficient knowledge and skills to work with the target group? |
| Interdisciplinary collaboration | Collaborators | Who do you see as your primary collaborators in a sick leave process?<br>What is your role and experiences with collaborating with the person on sick leave, relatives, case managers, employers, and others?<br>When do you feel that the collaboration works well and less well?<br>What would you like more or less of in the collaboration? |
| Themes | Exercise 1 | Which things are particularly prominent for you? How? Why? |
| Vocational rehabilitation | Exercise 2 | Does practice live up to the ideal? Why/why not? |
| Wishes for the future | The future ideal sick leave system | What could be the goal of future employment-oriented rehabilitation?<br>Do you have any ideas on what we could do in a better way? |

### 11.1.3 INTERVIEW GUIDE FOR CASE WORKERS

#### Agenda

1. Information, participant data etc.
2. Establishing ground rules
3. Talking about the target group: Defining them and how to approach them
4. Exercise 1 and 2
5. Intersectoral and -disciplinary collaboration (Exercise 3)
6. Rounding up

#### Ground rules

1. You are the expert → Say it as YOU see it
2. It’s about different experiences → Contribute with your perspective
3. I disagree/doubt what we are talking about → Share it with the group
4. I really want to say something → Respect the speaking order
5. Can I really say that? YES, honesty above all
6. I think a topic/aspect of discussion is missing → Say it out loud
7. Other??

#### The target group

People on sick leave due to common mental disorders have a higher-than-average risk of long-term sick leave, commonly referred to as being “vulnerable”.

1. What makes people vulnerable? Is it the people themselves or factors around them?
2. How much do they hold? How often do you deal with them?
3. Are you doing something different with vulnerable individuals with mental illness?
4. Is the system well set up for you to help them? How?
5. Are you missing something to better help the target group?
6. Other??

**Exercise 1: Discuss how you relate to the suggested themes**

**Exercise 2: Discuss the ideal of vocational rehabilitation**

**Exercise 3: Place the main actors in the matrix and explain why**

1. Add any missing actors
2. Associate the words that characterize each square/actor
3. Are there too many actors? Are they the right actors?
4. What would it look like in the ideal world?
5. Could a social medicine medical adviser make a difference? How?

## 11.2 INTERVIEW EXERCISES

### 11.2.1 EXERCISE 1: CONCEPTUAL PAIRS CONCERNING COLLABORATION IN VOCATIONAL REHABILITATION

Trust / Distrust

Security / Insecurity

Cohesion / Division

Order / Chaos

Cooperation / Counteroperation

Support / Resistance

Predictable / Unpredictable

Meaningful / Meaningless

Week / Day

Flexibility / Rigidity

Inclusive/Exclusive

Vulnerability / Robustness

Constructive / Destructive

Logical / Illogical

Possibilities / Limitations

Activating / Paralyzing

Manageable / Unmanageable

Targeted / Unfocused

Hopeful / Despondent

Person focus / System focus

??

### 11.2.2 EXERCISE 2 – THE IDEAL OF VOCATIONAL REHABILITATION

#### Rehabilitation

Rehabilitation is targeted **people**, which **experience or are at risk of experiencing limitations** in their physical, **psychological, cognitive and/or social functioning** and thus i **everyday life**.

The purpose of rehabilitation is a **meaningful life** with the best possible **activity and participation, coping and quality of life**.

Rehabilitation is one **collaborative process** between one **person, relatives, professionals and other relevant parties**.

**Rehabilitation efforts** is **targeted, coherent and knowledge-based** with **starting from the person’s perspectives and entire life situation**.

#### Employment-oriented rehabilitation

**Persons of working age** with **health-related impairments and the following reduced ability to work**

**The aim is to optimize work participation** through **different activities delivered by different actors in different settings**.

**How do the two definitions fit together? How can they not?**

### 11.2.3 EXERCISE 3: MAPPING STAKEHOLDERS INVOLVED IN VOCATIONAL REHABILITATION

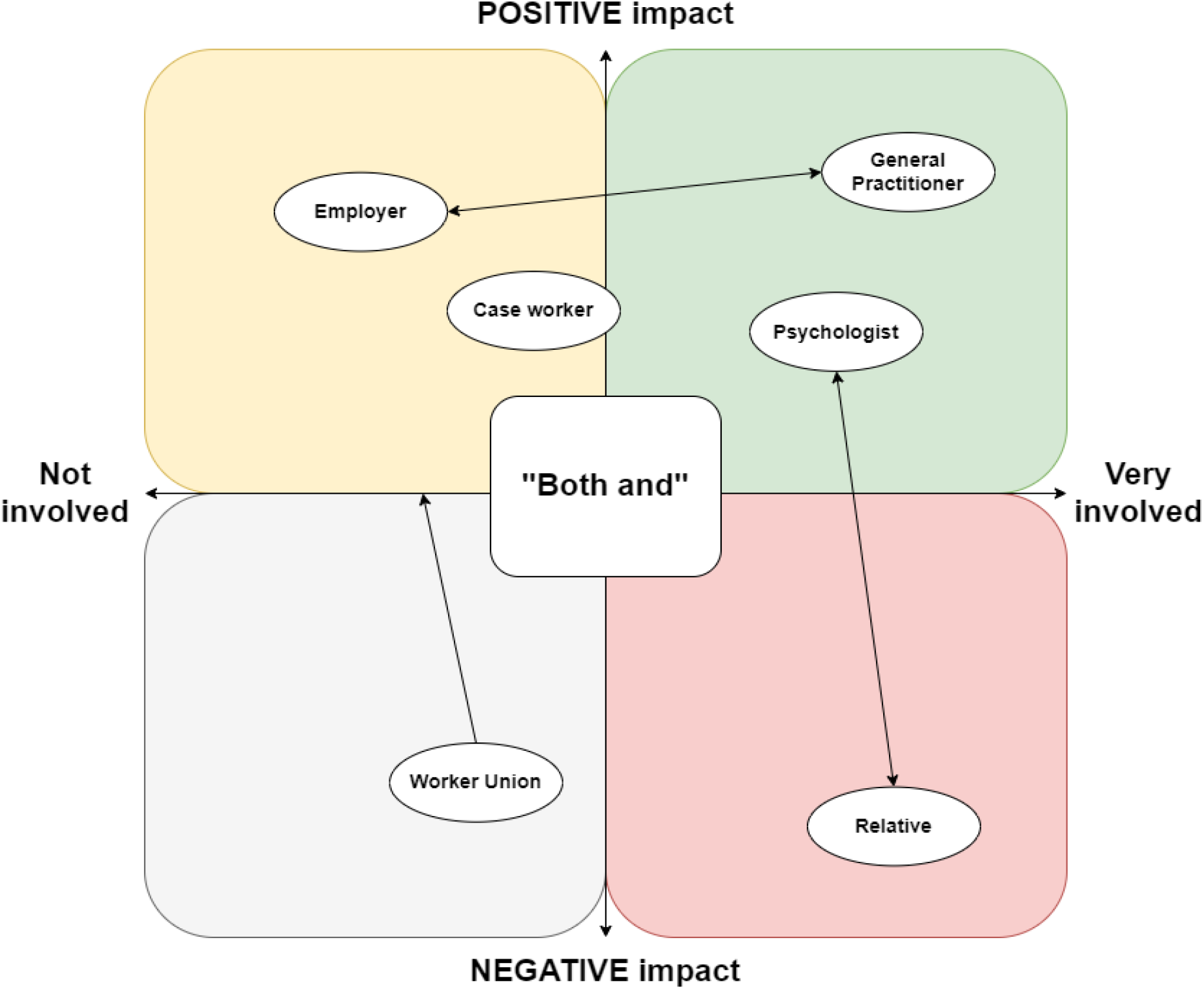

0.1 **A 2-dimensionla matrix about how the person on sick leave being interviewed has experienced the involvement of stakeholders in his/her sick leave process. The person being interviewed was asked to rate how much each type of stakeholder, e.g**., **the general practitioner, had been involved (the horizontal axis) and whether this had had a positive or negative impact on the process (the vertical axis). There were about 15 common stakeholders in vocational rehabilitation that the interviewee could choose from and 4 blank “puzzles” that could be filled in during the interview if the interviewee felt that anyone was missing from the list. The arrows indicate how people described that the role of stakeholders changed over the course of the sick leave process. Arrows are examples and not general findings**.

## 12 TABLES

### 12.1 TABLE 1A-1C

**Table 1a.** Characteristics of citizens.

| Variable | municipality 1 | municipality 2 | Total |
| --- | --- | --- | --- |
| Sex (Female/Male) | 2/2 | 3/0 | 5/2 |
| Age, range (years) | 45-61 | 35-54 | 35-61 |
| Partner (yes/no) | 1/3 | 2/1 | 3/4 |
| Job status (sick leave/partial sick leave/laid off) | 0/1/3 | 0/0/3 | 0/1/6 |
| Sick leave duration, range (days) | 113-209 | 195-1645 | 113-1645 |
| Sick leave duration, median (days) | 197 | 807 | 206 |
| Job experience, range (years) | 12-40 | 9-31 | 9-40 |
| Job experience, median (years) | 36 | 10 | 31 |
| Former sick leave, range (N) | 1-10 | 1-7 | 1-10 |
| Former sick leave, average (N) | 5 | 4 | 5 |
| Physical comorbidities (yes/no) | 2/2 | 0/4 | 2/6 |
| Mental comorbidities (yes/no) | 1/3 | 1/2 | 2/5 |
| Former Job | Kindergarden teacher, Accounting assistant, High School secretary, Bar and shop manager | Case worker in unemployment insurance fund, On-call substitute at a shelter, Occupational Therapist in a rehabilitation centre |  |
| Education | Pedagogue. Accounting, Secretary, 10th grade | Administration, painter, general upper secondary education |  |

**Table 1b.** Characteristics of CWs.

|  | municipality 1 |  |  | municipality 2 |  |  | Total |
| --- | --- | --- | --- | --- | --- | --- | --- |
| Variable | EXP | INE | MIX | EXP | INE | JRC |  |
| Participants, N | 5 | 4 | 6 | 6 | 3 | 7 | 31 |
| Age, range (years) | 53-64 | 22-30 | 25-63 | 42-65 | 25-30 | 36-54 | 22-65 |
| Age, median (years) | 56 | 29 | 43 | 49 | 29 | 50 | 46 |
| Job experience, range (years) | 13-36 | 0-6 | 1-45 | 7-42 | 2-7 | 9-25 | 0-45 |
| Job experience, median (years) | 20 | 3 | 7 | 15 | 6 | 13 | 13 |

All CWs were female.
Educational background of CWs: Of the 31 CWs, 22 (71%) were held a bachelor's degree in social work (professional degree), while 9 CWs (29%) had other educational backgrounds, including a master's degree in Economics and Business Administration, a master's degree in Coaching, a bachelor's degree in Education, a master's degree in Sustainable Cultural Heritage Management, a master's degree in Political Science, a master's degree in Psychology and Communication, and a master's degree in Business Language and Communication. EXP Experienced CWs, INE Inexperienced CWs, MIX Mixed group of CWs working exclusively with long-term sick leave cases, JRC Job retention specialists

**Table 1c.** Characteristics of general practitioners.

| Variable | municipality 1 | municipality 2 | Total |
| --- | --- | --- | --- |
| Sex (Female/Male) | 2/1 | 2/2 | 4/3 |
| Age, range (years) | 49-60 | 44-62 | 44-62 |
| Job experience, range (years) | 8-23 | 5-21 | 5-23 |
| Job experience, median (years) | 23 | 10 | 10 |
| Organization (Partnership/solo practice) | 3/0 | 2/2 | 5/2 |
| Patients in practice, range (N) | 3300-3883 | 1620-3300 | 1620-3883 |
| Patients in practice, median (N) | 3400 | 3250 | 3300 |

#### Non-participation

Six citizens originally interested in the study when approached by the first author ended up not participating in interviews (46%). Most citizens did not respond following initial contact when the first author tried to schedule interviews, and others replied that they did not having the energy. We contacted 16 GPs, of these 7 participated (44%). Most GPs simply did not get back to use after the first author had informed about the study, most commonly through the GP’s secretaries, and others declined due to lack of time. Focus groups with CWs included a total of 31 CW, corresponding to about 80-85% of the total number of CWs in the sickness benefits department. Those that did not attend did not provide reasons as this was not demanded.

### 12.2 TABLE 2

**Table 2.** Means and ends in vocational rehabilitation.

| Actor | Principle/Logic | Means | End (Functional goal) |
| --- | --- | --- | --- |
| Citizen | "Access to care should precede participation in return-to-work activities" | Access to care-related activities is regarded as a premise for being able to take part in return-to-work activities. | Improved functioning leading to return to work to the extent possible. |
| CW | "Focusing on people's potential to return-to-work improves the value of care activities" | Workfare principles are used to steer the focus in care towards improving people's potential to return to work. | Motivation and readiness for work-related activities during care |
| GP | "Focusing on people's health care needs in return-to-work activities improves value of return-to-work activities" | Advocating for the interrelationship between health and work by emphasizing the work benefits of health services and vice versa the health benefits of work services. | Consciousness about implications of citizens' health on work-related activities and vice versa. |

### 12.3 TABLE 3

**Table 3.** Definition of theoretical concepts in themes.

| Concept | Definition and sources |
| --- | --- |
| Shared goals | *Shared goals concerns where people have mutual goals to achieve in agreement, which motivate them to move beyond **subgoal optimization** to act with greater regard for the whole, thus transcending their specific **functional goals** (53,83). The opposite of shared goals is functional goals or subgoal optimization where tasks are solved based on specialized work goals without reference to the overarching goals of the work process (83,84).* |
| Shared knowledge | *Shared knowledge concerns the level of knowledge collaborators have about each other's training, expertise, and role, which enables them to see how their specific tasks interrelate with the whole process, thus facilitating systems thinking about how their tasks and the tasks of others contribute to the whole (53,85,86). The opposite of shared knowledge is specialized knowledge where tasks are solved without having shared knowledge about each other (84).* |
| Mutual trust and respect | *Mutual respect concerns having respect for the work of others, i.e., recognizing each person's pride and status related to their role to overcome potential status barriers that prevent them from seeing and taking account of the work of others, to value their contributions, and to consider the impact of one's actions on others, thus acting with respect for the overall work process (53,83,87,88). The opposite of mutual respect is disrespect where the work of others is disrespected (84).*<br>*Trust can be viewed from an individual's perspective as a psychological state comprising the intention to accept vulnerability based on positive expectations of the intentions or the behavior of another. From a more sociological viewpoint, trust is viewed less as a characteristic of an individual, and more as an attribute of a social relationship between two or more actors. From this relational perspective, trust is defined as existing when one party to the relation* |
|  | *believes the other party has incentive to act in his or her interest or to take his or her interest to heart (89).*<br>*Trust concerns **people’s beliefs about the intentions of others.** Respect concerns **the degree to which people recognize the work of others and adapt their behaviour accordingly.** From this distinction, two distinct aims for the relationship between parties emerge: An ambition of trustworthiness, e.g., being trustworthy yourself or feeling that others are worth trusting, and an ambition of mutual respect, i.e., feeling that others value and take account of your contribution or trying to value and take account in your behaviour of other people’s contributions. Both respect and trust can be mutual, i.e., coming from both parties of a relation, or unilateral, i.e., only coming from one of the parties of a relation. We view the relationship between trust and respect as bidirectional, i.e., they affect each other and can both result from the other.* |

## 13 FIGURES (TIFF, POSTSCRIPT OR EPS FILES)

### List of figures

1. Figure 1 Caption: Collaborative Challenges in Vocational Rehabilitation Due to Weak Relational Ties, Negative Collaborative Experiences, and Counterproductive Communication Strategies: An explanatory framework
2. Figure 1 Alt Text: An explanatory framework illustrating how weak relational ties sustain collaborative challenges in vocational rehabilitation. On the left, a blue circle labelled “Weak Relational Ties” contains three elements: lack of shared goals, limited shared knowledge, and low mutual trust and respect. In the centre, a red circle labelled “Negative Collaborative Experiences” contains three experiences: competing for the agenda, finding collaboration “colossally labouring,” and perceiving collaborators as obstacles. On the right, a yellow circle labelled “Counterproductive Communication Strategies” contains three communication strategies: campaigning, clarifying, and self-protective communication. Arrows connect the three circles in a continuous cycle, illustrating how weak relational ties give rise to negative collaborative experiences, which participants attempt to navigate through communication strategies that often reinforce the underlying relational challenges.

