## Supplementary material for "Weak relational ties and ineffective communication in vocational rehabilitation: a case study of people on long-term sick leave with common mental disorders": Implications for rehabilitation

*2-4 points*

*A feature of this journal is a boxed insert on Implications for Rehabilitation. This should include between two to four main bullet points drawing out the implications for rehabilitation for your paper. This should be uploaded as a separate document.*

1) Collaboration in vocational rehabilitation for people on long-term sick leave due to common mental disorders may be improved by strengthening relational coordination among stakeholders.
2) Strengthening relational coordination in vocational rehabilitation will require careful consideration about fostering shared goals, shared knowledge, mutual trust and respect, and high-quality communication across professional and organizational boundaries.
3) High-quality communication among stakeholders in vocational rehabilitation is key to prevent counterproductive communication strategies from sustaining negative relational dynamics.
4) Efforts to strengthen relational coordination include supportive organisational and policy conditions to manage workload, time constraints, and possibilities for coordination among stakeholders. Without addressing these conditions, initiatives targeting individual practices alone are unlikely to be effective and sustainable.
